# Circulating protein profiling identifies prognostic biomarkers in amyotrophic lateral sclerosis

**DOI:** 10.64898/2026.07.20.26354798

**Authors:** Hadar Klimovski, Marcel Weinreich, Andrew Strange, Iddo Magen, Nancy S Yacovzada, Yahel Cohen, Dganit Melamed Kadosh, David G Lester, Avigail Taylor, Yuhan Zhou, Cory Holdom, Tamar Ziv, Beatrice Abramovich, Anand G Subramaniam, Eran Perlson, Vivian Drory, Arie Admon, Pamela J Shaw, Andrea Malaspina, Martin R Turner, Kevin Talbot, Alexander G Thompson, Johnathan Cooper-Knock, Eran Hornstein

## Abstract

Amyotrophic lateral sclerosis (ALS) is a fatal neurodegenerative disease characterized by progressive motor neuron loss and marked clinical variability. Blood-based biomarkers offer a scalable approach to stratify patients and support therapeutic development. In a large-scale serum proteomic survival study of 730 patients with ALS from independent cohorts in Israel and the UK, we used complementary proteomic technologies, mass spectrometry and the Olink proximity extension assay (PEA), enabling cross-cohort and cross-platform evaluation of concordant protein effects. We identified serum proteins associated with survival in both cohorts. GSN, IGFBP2 and MEGF10 added prognostic information beyond clinical variables and neurofilament light chain (NfL). These circulating proteins are therefore candidate biomarkers for improved prognostic assessment that may lead to improved precision-medicine approaches in ALS.

## Introduction

Amyotrophic lateral sclerosis (ALS) is a fatal, progressive neurodegenerative syndrome marked by selective loss of upper and lower motor neurons in the brain and spinal cord, with a median survival of 30 months from onset of weakness. Marked clinical heterogeneity in survival and disease progression rates^1^ complicates counseling and trial design, underscoring the need for robust biomarkers to improve patient stratification^2,3^, disease management and therapeutic development.

Blood-based molecular profiling increasingly captures signatures of tissue damage, stress and immune activation in neurodegenerative diseases such as ALS^4^, and neurofilament light chain (NfL) is a validated, robust prognostic biomarker^2,3^. However, most other candidate biomarkers have failed to reach clinical use owing to poor reproducibility and insufficient validation, and systematic analyses of circulating serum proteins for ALS prognostication remain limited^5–7^.

To address this gap, we applied an unbiased proteome-wide approach to two large independent ALS cohorts, from Israel (n = 350) and the UK (n = 380), profiled by mass spectrometry (1,218 proteins) and the Olink PEA (5,416 proteins), respectively. Time-to-event analysis of the 707 proteins shared across platforms identified three proteins that add prognostic value beyond established clinical covariates.

## Results

### Serum proteins linked to ALS survival

To identify candidate prognostic biomarkers for ALS, we profiled demographically diverse cohorts from Israel (n = 350) and the UK (n = 380) with a cross-platform approach integrating mass spectrometry and the Olink PEA, and evaluated 707 shared proteins in time-to-event analyses. The study design and patient characteristics are summarized in Fig. 1 and Table 1.

**Fig 1.**
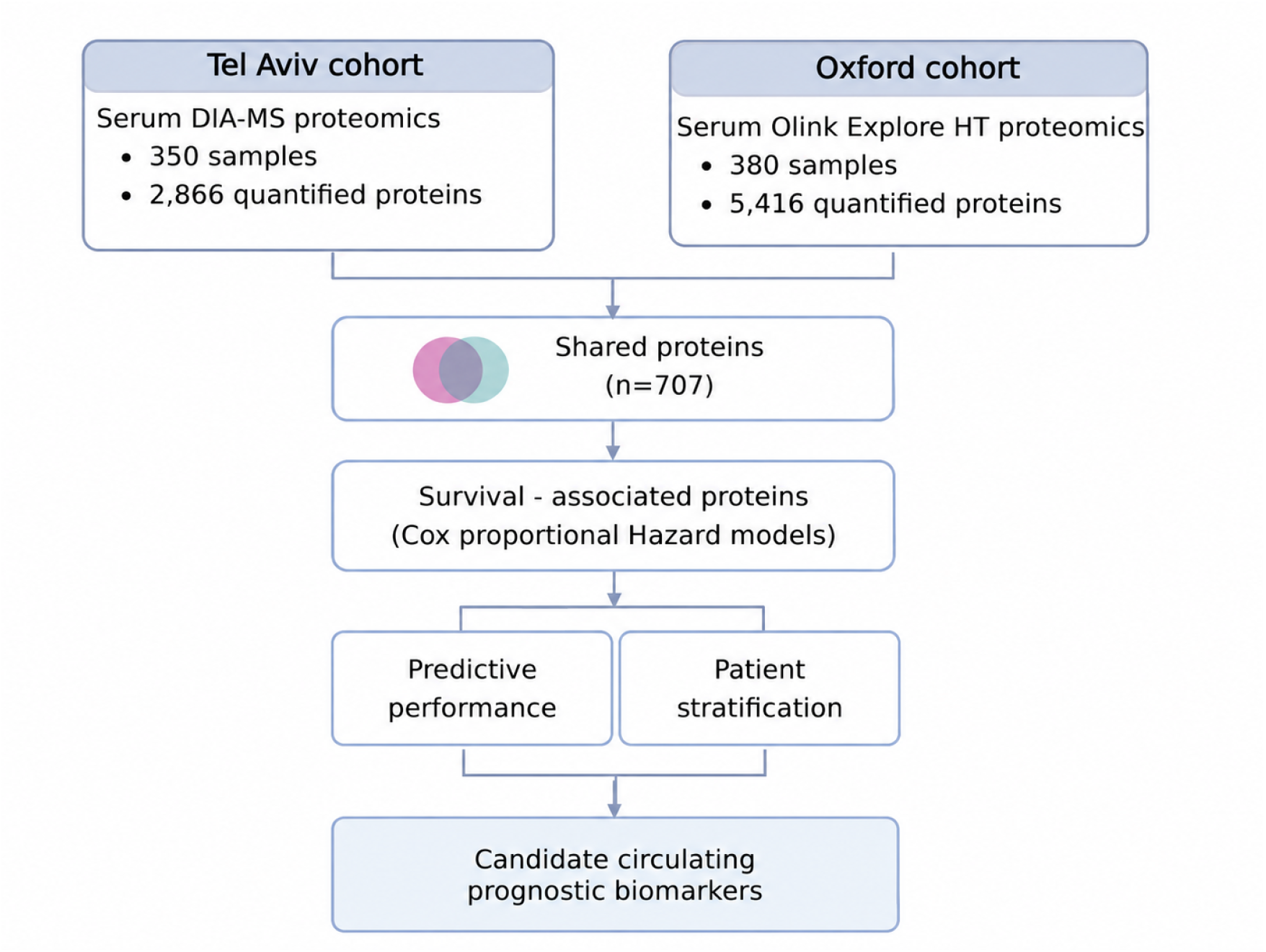
Study design and analytical workflow for cross-cohort validation of serum proteomic survival biomarkers in ALS. The Tel Aviv cohort (n = 350) was profiled using data-independent acquisition mass spectrometry (DIA-MS), yielding 2,866 and 1,218 proteins before and after quality control, respectively. The Oxford cohort (n = 380) was profiled with the Olink PEA, yielding 5,416 proteins. A total of 707 overlapping proteins were evaluated using either unadjusted or demographic-adjusted Cox proportional hazards models, and proteins with significant survival associations in both cohorts (FDR < 0.05 after adjustment) were taken forward for prognostic modelling and risk-group analysis.

**Table 1.** Clinical characteristics of patients with ALS at serum collection. Non-normally distributed variables are reported as medians with interquartile ranges (Q1–Q3), normally distributed variables as means ± s.d. Categorical variables as counts (n) and percentages (%). Median survival from symptom onset was estimated using the Kaplan–Meier method. Higher forced vital capacity (FVC) values indicate better pulmonary capacity. Genetic testing results are provided for pathogenic *C9orf72* or SOD*1* variants. Bulbar and limb onset were classified by the initial site of symptom emergence. Survival from symptom onset (years) was defined as the interval between the first reported ALS symptoms and either death or censoring.

Serum from the Tel Aviv cohort (350 patients with ALS recruited at Tel Aviv Sourasky Medical Center) was profiled by data-independent acquisition mass spectrometry (DIA-MS), quantifying 2,866 proteins. After quality control, which retained proteins present in ≥50% of patients and excluded those with low variability or high correlation, 1,218 proteins remained for analysis.

To evaluate reproducibility across cohorts and proteomic technologies, we analyzed an independent UK cohort of 380 patients with ALS recruited at the John Radcliffe Hospital, Oxford, whose serum was profiled with the affinity-based Olink PEA, quantifying 5,416 proteins^8^. In total, 707 proteins were shared between the Tel Aviv DIA-MS and Oxford Olink datasets and were taken forward for analysis.

Associations between protein abundance and survival were assessed using univariate Cox proportional hazards models.

Fifteen of the 707 shared proteins were significantly associated with survival in both cohorts (FDR < 0.05; **Fig. 2a,b**), with strong cross-cohort concordance of their effect sizes (Pearson R = 0.96, P = 2 × 10⁻⁸; **Fig. 2c**). These included tumor necrosis factor receptor superfamily member 12A (TNFRSF12A), insulin-like growth factor-binding protein 2 (IGFBP2), inter-alpha-trypsin inhibitor heavy chain 3 (ITIH3), growth hormone receptor (GHR) and gelsolin (GSN; Extended data table 1). Directional concordance extended to all 707 shared proteins (Pearson R = 0.46, P = 3.6 × 10⁻³⁸; **Extended Data Fig. 1a**), indicating consistent survival-associated proteomic patterns across independent cohorts and orthogonal platforms.

**Figure 2.**
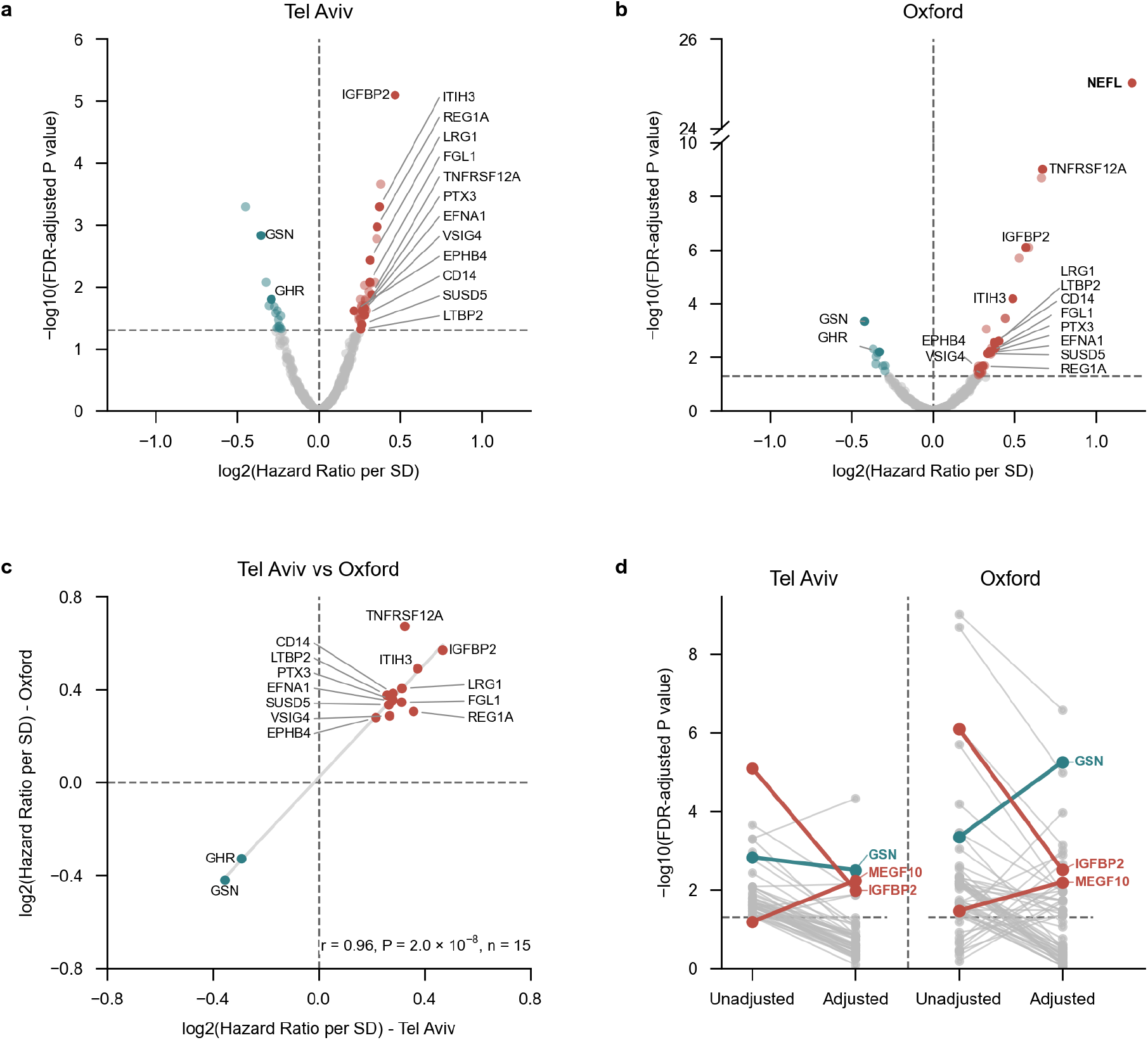
Cross-cohort ALS survival-associated proteomics. **a,b,** Volcano plots of HRs from univariate Cox proportional hazards regression in the Tel Aviv (**a**, *n* = 350) and Oxford (**b**, *n* = 380) cohorts, showing log₂-transformed HRs (*x* axis) versus −log₁₀ FDR-adjusted *P* values (*y* axis). Proteins with FDR < 0.05 are coloured by direction of association (teal, protective; red, risk); grey indicates non-significant proteins. The dashed horizontal line marks FDR = 0.05. **c,** Cross-cohort Pearson correlation of log₂-transformed HRs for the 15 proteins significantly associated with survival in both cohorts (FDR < 0.05). **d,** Slope chart showing the significance of Cox models before and after adjustment for age at symptom onset and sex. Colour denotes direction of effect (red, HR > 1; blue, HR < 1), and the dashed horizontal line marks FDR = 0.05.

### Age and sex reshape survival-associated proteomic signals

Age at symptom onset and biological sex are established modifiers of disease course and survival^9,10^ and are associated with changes in the serum proteome. Adjustment for these covariates changed the significance of associations across shared proteins (Fig. 2d). In the Tel Aviv cohort, only eight of 48 survival-associated proteins remained significant after adjustment (FDR < 0.05), as did 28 of 47 in the Oxford cohort (Extended Data Fig. 1b–e; Extended data table 1), suggesting that age and sex account for part of the survival-associated proteomic signal. Notably, GSN, IGFBP2 and MEGF10 remained significantly associated with survival in both cohorts after adjustment for age at symptom onset and sex and correction for multiple testing (FDR < 0.05). Higher GSN levels were associated with longer survival in both cohorts (Tel Aviv: hazard ratio (HR) per standard deviation (s.d.) = 0.77, 95% CI 0.69–0.87, P = 8.85 × 10⁻⁶; Oxford: HR = 0.70, 95% CI 0.62–0.79, P = 1.58 × 10⁻⁸), whereas higher IGFBP2 and MEGF10 levels were associated with shorter survival (Tel Aviv: IGFBP2, HR = 1.27, 95% CI 1.13–1.43, P = 8.83 × 10⁻⁵; MEGF10, HR = 1.29, 95% CI 1.15–1.46, P = 2.45 × 10⁻⁵; Oxford: IGFBP2, HR = 1.36, 95% CI 1.18–1.58, P = 3.92 × 10⁻⁵; MEGF10, HR = 1.27, 95% CI 1.13–1.44, P = 9.91 × 10⁻⁵). Other proteins showed cohort-specific survival associations, including TNFRSF12A, PTX3, IGFBP6 and ART3 (Oxford only) and GRN, PEBP4 and IL1RAP (Tel Aviv only).

In a hypothesis-generating analysis of Human Protein Atlas single-cell RNA profiles, *GSN* and *IGFBP2* were broadly expressed across peripheral cell types, whereas *MEGF10* was preferentially enriched in glial cells and skeletal muscle satellite cells (**Extended Data Fig. 2**).

### Bootstrap selection identifies reproducible prognostic proteins across cohorts

As an orthogonal test of robustness, we used bootstrap stability selection. For each cohort, we fitted Cox proportional hazards models adjusted for age at symptom onset and sex across 1,000 bootstrap iterations, and defined selection frequency as the proportion of iterations in which a protein reached significance after within-iteration false discovery rate correction (FDR < 0.05).

To distinguish stable associations from chance, we set cohort-specific thresholds at the 95th percentile of the maximum selection frequency across permutations, which capped the probability of chance selection at ∼5% (83.0% in Tel Aviv and 76.0% in Oxford; Fig. 3a,b). Six proteins in Tel Aviv and 17 in Oxford exceeded these thresholds. Three proteins, GSN, IGFBP2 and MEGF10, were selected in both cohorts (Fig. 3c). Bootstrap stability selection therefore corroborated the primary Cox regression findings, independently identifying GSN, IGFBP2 and MEGF10 as robust survival-associated proteins in both cohorts.

**Figure 3.**
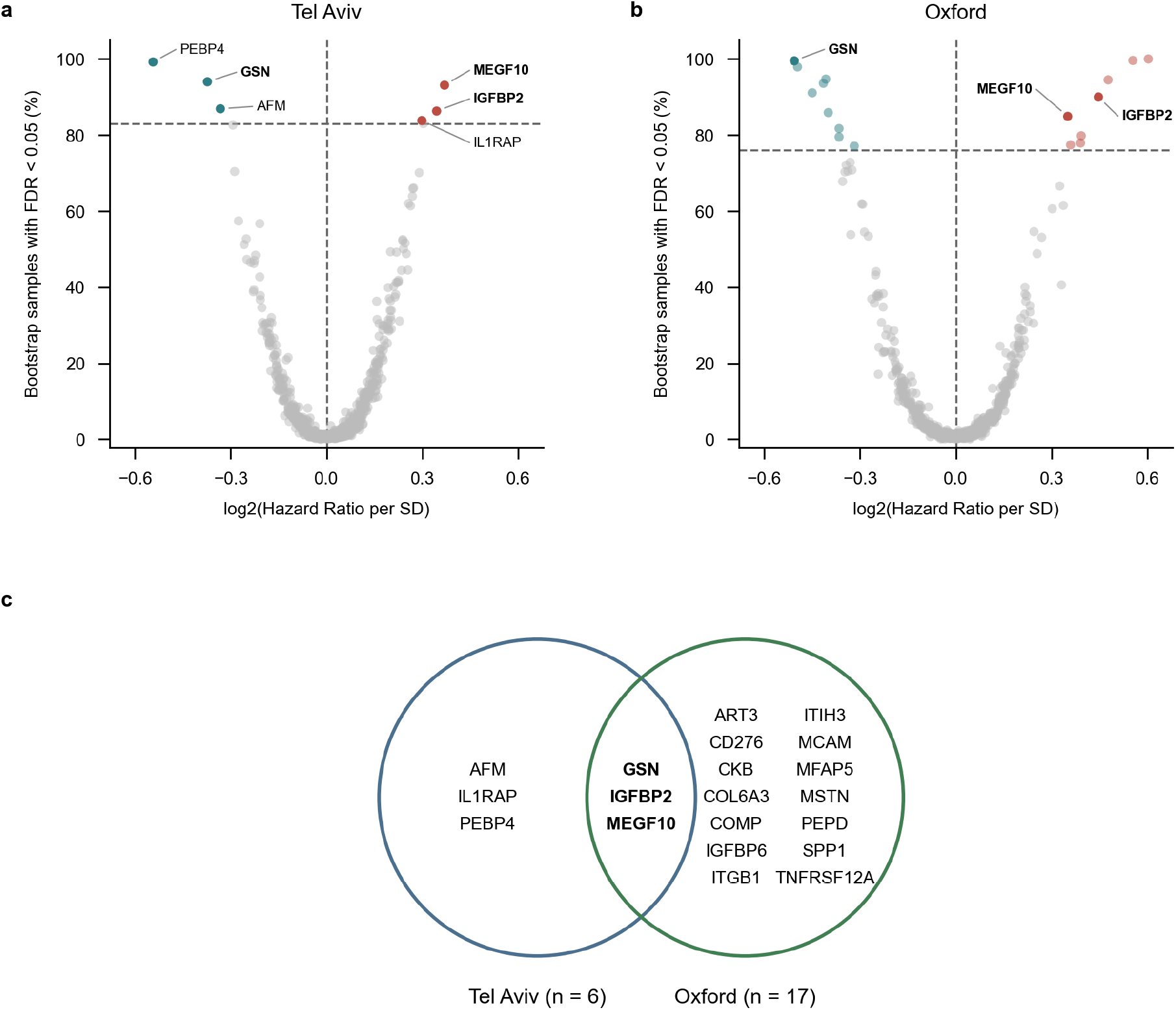
Bootstrap stability selection of candidate prognostic proteins across independent ALS cohorts. **a,b,** Volcano plots of bootstrap percentage selection frequency at FDR < 0.05 (*y* axis) plotted against log₂-transformed HR per s.d. increase in protein abundance (*x* axis) in the Tel Aviv (**a**) and Oxford (**b**) cohorts, adjusted for age at symptom onset and sex. Horizontal dashed lines indicate permutation-derived selection thresholds (83% and 76%, respectively). Proteins exceeding the thresholds are coloured by direction of association (red, HR > 1; blue, HR < 1), whereas proteins below the thresholds are shown in grey. **c,** Venn diagram showing the overlap of proteins exceeding the selection thresholds in the Tel Aviv and Oxford cohorts.

### GSN, IGFBP2 and MEGF10 add prognostic value in ALS

To determine whether GSN, IGFBP2 and MEGF10 add prognostic value beyond established clinical predictors, we fitted a flexible parametric (Royston–Parmar) survival model, with time measured from blood sampling, and evaluated it by five repeats of 20-fold cross-validation. The clinical model included age at symptom onset, sex, site of onset, time from symptom onset to sampling and ALSFRS-R slope, together with forced vital capacity (FVC) and *C9orf72* status. Adding GSN, IGFBP2 and MEGF10 improved prognostic accuracy in both cohorts (**Fig. 4a**): ΔC-index (Tel Aviv +0.026, 95% CI 0.000–0.050; Oxford +0.059, 95% CI 0.031–0.088), 2-year ROC ΔAUC (Tel Aviv +0.044, 95% CI 0.003–0.083; Oxford +0.06, 95% CI 0.015–0.103) and the integrated Brier score (ΔIBS, Tel Aviv 0.01, 95% CI 0.001–0.019; Oxford 0.02, 95% CI 0.01–0.03). Each protein also added prognostic value individually (**Fig. 4a** and **Extended data table 4**).

**Fig 4.**
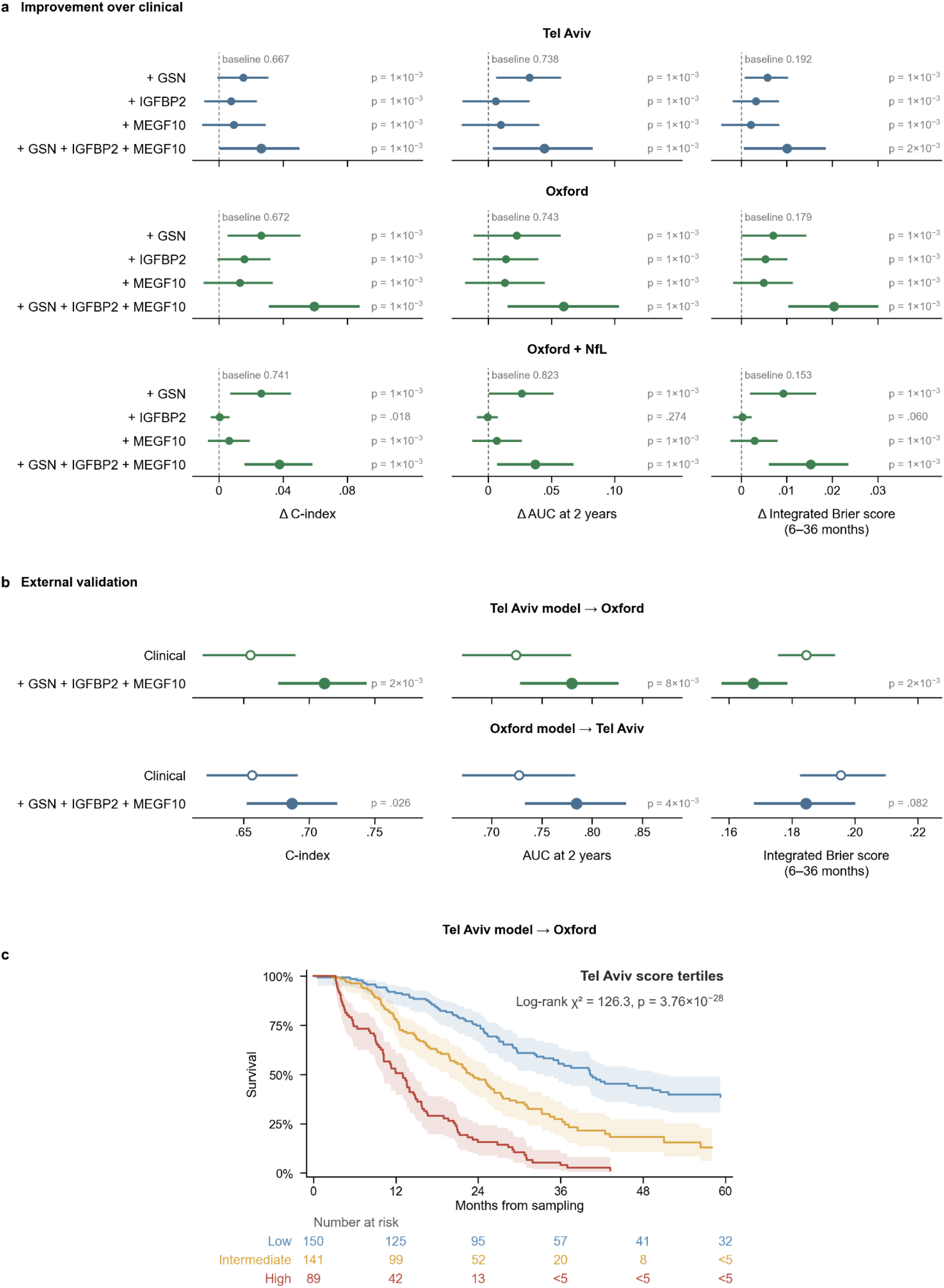
GSN, IGFBP2 and MEGF10 add prognostic information beyond clinical covariates and NfL in ALS. **a,** Incremental predictive performance of GSN, IGFBP2 and MEGF10, individually and in combination, over the clinical model in the Tel Aviv (n = 350) and Oxford (n = 380) cohorts and over the clinical model plus serum neurofilament light chain (NfL) in Oxford, assessed by C-index, time-dependent area under the curve (AUC) at 2 years and integrated Brier score (6–36 months). Positive values indicate improvement over the baseline model; circles and error bars denote gains from pooled out-of-fold predictions with 95% bootstrap CIs. **b,** Cross-cohort external validation of a locked model combining clinical features with GSN, IGFBP2 and MEGF10 without refitting, using standardized protein abundances and paired bootstrap P values. **c,** Kaplan–Meier survival curves in the Oxford cohort stratified by risk-score tertile thresholds derived from the Tel Aviv three-protein model. log-rank χ² = 126.3, P = 3.76 × 10⁻28.

We further evaluated the incremental value of GSN, IGFBP2 and MEGF10 beyond a clinical reference model that included the established prognostic biomarker NfL^3,11^, which was measured in the Oxford cohort. The three proteins improved discrimination (ΔC = +0.038, 95% CI 0.016–0.058), increased the 2-year AUC (ΔAUC = +0.037, 95% CI 0.007–0.068) and reduced the integrated Brier score (ΔIBS = 0.015, 95% CI 0.006–0.024; permutation *P* < 0.001 for all three metrics).

To evaluate generalizability, we trained a model combining GSN, IGFBP2, MEGF10 and clinical features on one full cohort and applied it to the other cohort without refitting or using its outcomes; validation-cohort protein abundances were standardized to account for scale differences between DIA-MS and Olink (Fig. 4b). Adding the three proteins improved discrimination in both training-to-validation directions (Tel Aviv to Oxford: ΔC = +0.057, *P* < 0.001; Oxford to Tel Aviv: ΔC = +0.030, *P* = 0.024), with consistent improvements in 2-year AUC (ΔAUC = +0.056, *P* = 0.006 and +0.057, *P* = 0.002, respectively). The integrated Brier score decreased in both directions, significantly only in Oxford (ΔIBS = 0.017, *P* < 0.001 and ΔIBS = 0.011, *P* = 0.080, respectively; **Fig. 4b** and **Extended Data Table 4**). Cross-cohort calibration is shown in **Supplementary Fig. 3b**.

Finally, risk-score tertile cut-offs derived from the Tel Aviv three-protein model separated Oxford patients into low-, intermediate- and high-risk groups (n = 150, 141 and 89) with median survival from sampling of 40.4, 23.2 and 13.0 months, respectively (Kaplan–Meier analysis; log-rank χ² = 126.3, P = 3.76 × 10⁻^28^; Fig. 4c).

### Cross-cohort pathway analysis and pre-diagnostic protein dynamics

Gene-set enrichment analysis of 1,289 Tel Aviv and 5,410 Oxford proteins ranked by signed Cox Wald statistics revealed concordant positive enrichment of immune-related pathways across cohorts, including inflammatory response regulation, cytokine signaling, innate immune activation and neutrophil migration (**Fig. 5a**). Immune pathways were significantly more likely than non-immune pathways to show positive enrichment in Oxford (odds ratio [OR] = 2.7, P = 3 × 10⁻⁷) and Tel Aviv (OR = 1.9, P = 3 × 10⁻³; Fig. 5a,b), indicating cross-cohort convergence of survival-associated proteomic signals on immune and inflammatory processes. Of 1,765 pathways tested in both cohorts, 20 reached nominal significance (P < 0.05) in both, with concordant enrichment directions. These included pathways related to muscle contraction NF-κB signalling, apoptotic signalling and phagocytosis recognition (Fig. 5c).

**Fig 5.**
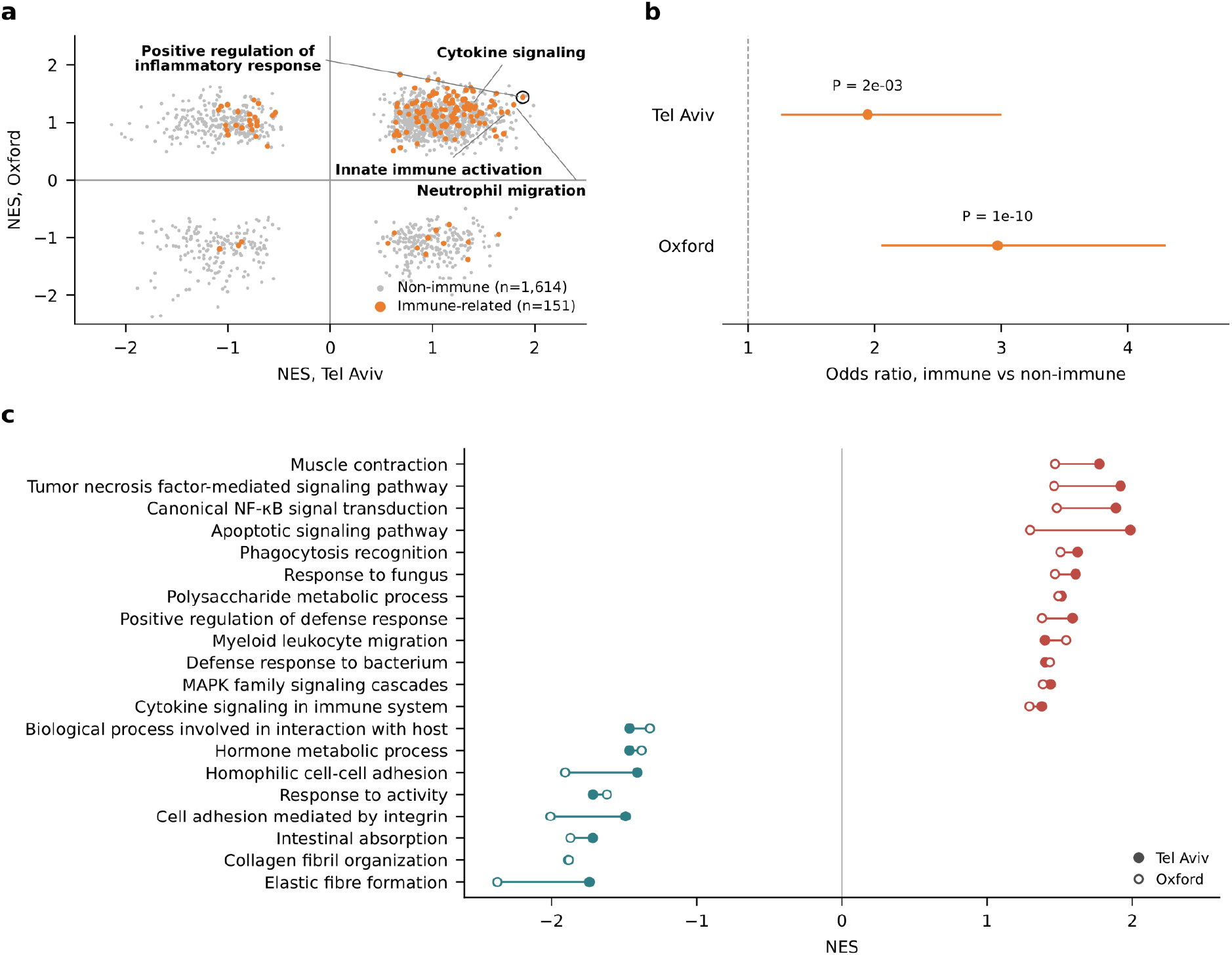
Proteomic pathway enrichment. **a,** Comparison of pathway normalized enrichment scores (NES) between the Tel Aviv and Oxford cohorts (immune pathways, brown; non-immune pathways, grey). **b,** ORs and 95% CI for immune-related versus non-immune pathways per cohort. Two-sided Fisher’s exact test *P* values. **c,** 20 shared pathways across cohorts (concordant NES direction at *P* < 0.05; red, positive NES; teal, negative NES). Full results are in Extended data Table 5.

To examine pre-diagnostic dynamics, we analysed UK Biobank^12^ plasma proteomic profiles of GSN, IGFBP2, MEGF10 and NEFL in participants with motor neuron disease (MND), all-cause dementia, Alzheimer’s disease or Parkinson’s disease and in healthy controls. All four proteins varied across groups, with distinct MND-enriched patterns (**Fig. 6a**): compared with controls (n = 4,567), individuals with MND (n = 281) had lower GSN (−0.21 s.d., FDR = 1.7 × 10⁻³) and elevated IGFBP2 (+0.19 s.d., FDR = 1.7 × 10⁻³), MEGF10 (+0.30 s.d., FDR = 4.7 × 10⁻⁴) and NEFL (+1.51 s.d., FDR = 2.2 × 10⁻³⁴).

**Fig 6.**
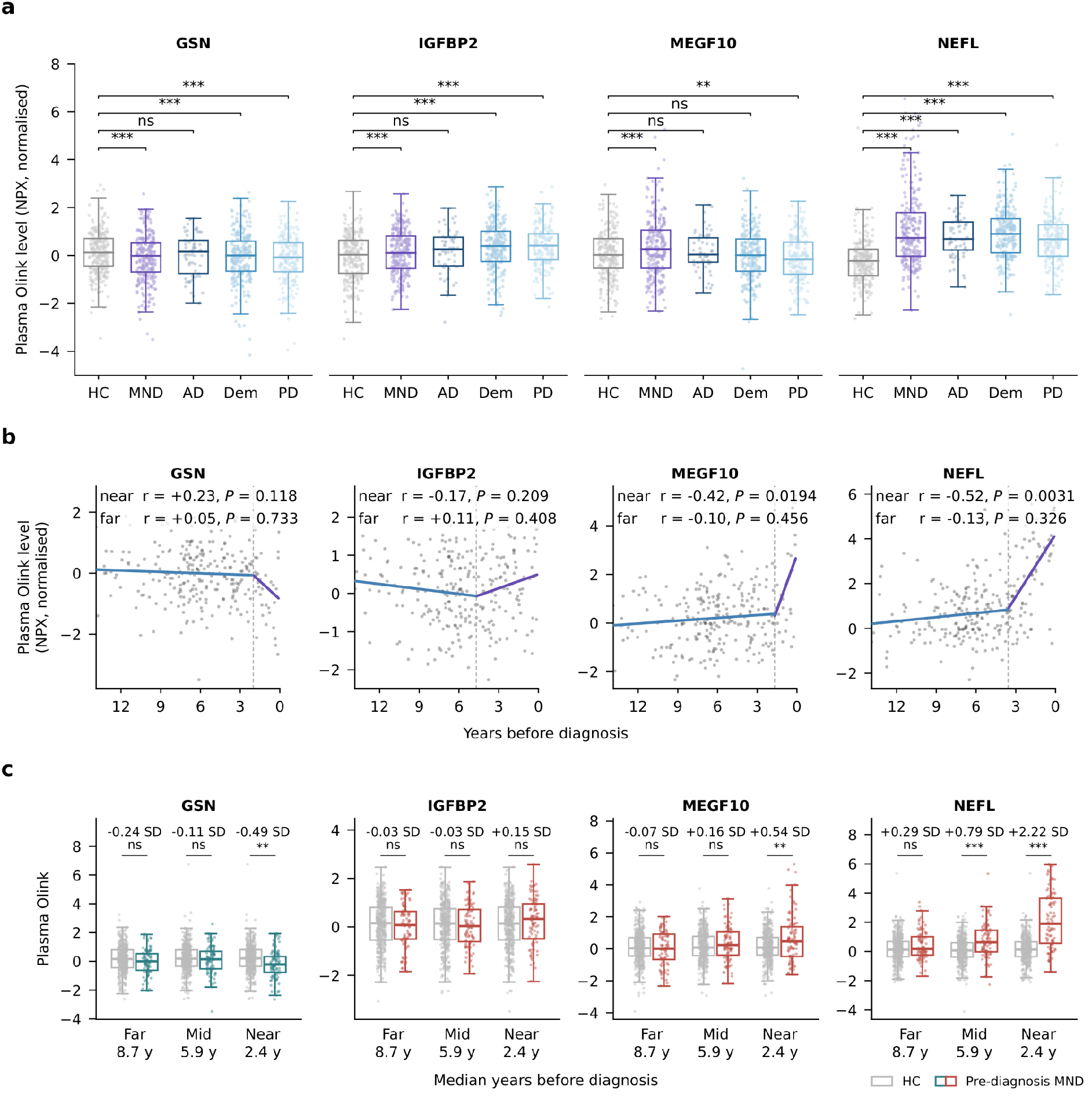
Pre-diagnosis plasma protein levels in UK Biobank. **a,** Plasma levels of GSN, IGFBP2, MEGF10 and NEFL in healthy controls and participants with ALS (motor neuron disease, MND), Alzheimer’s disease, all-cause dementia or Parkinson’s disease. Two-sided t-tests versus healthy controls (P < 0.05). **b,** Normalised protein levels versus time before diagnosis in pre-diagnostic ALS cases (MND, n = 259), with protein-specific inflection points estimated via fitted cubic smoothing splines . Correlations and two-sided P values are shown for intervals farther from (blue) and nearer to (purple) diagnosis. **c,** Protein levels in pre-diagnostic ALS (MND) cases versus age- and sex-matched healthy controls across far, intermediate and near tertiles of time before diagnosis. Effect sizes are shown in s.d. of matched controls. P values from two-sided t-tests of the case–control coefficient were adjusted across proteins tested within each tertile: ***P < 0.001, **P < 0.01, *P < 0.05; n.s., not significant. Colour denotes direction of effect (red, HR > 1; blue, HR < 1),

We next evaluated case–control differences across prespecified pre-diagnostic time windows (**Fig. 6b,c**). GSN, MEGF10 and NEFL differed significantly from matched controls approaching diagnosis, whereas IGFBP2 did not. In the window closest to diagnosis, GSN was reduced (−0.46 s.d., FDR = 4.1 × 10⁻³^8^) and MEGF10 elevated (+0.54 s.d., FDR = 4.1 × 10⁻³) in MND cases, and NEFL increased progressively towards diagnosis, reaching +0.79 s.d. in the intermediate window (FDR = 3.8 × 10⁻⁸) and +2.22 s.d. in the proximal window (FDR = 1.0 × 10⁻⁴¹).

Together with the work of Lester et al.^8^, these studies demonstrate the utility of high-throughput proteomics in large cohorts, clarify the advantages and limitations of cross-technology validation and identify candidate blood-based prognostic biomarkers for ALS.

## Discussion

The marked heterogeneity of ALS survival complicates clinical trial design and prognostic counseling. Although clinical baseline models and NfL measurements have substantially improved ALS prognostication, additional molecular biomarkers are needed. Circulating miRNAs have recently emerged as candidate prognostic biomarkers^13,14^, but the small number of validated molecular markers underscores the need for additional disease-relevant biomarkers with distinct or complementary sensitivity and specificity.

In this study, we analyzed serum proteomic data from 730 patients with ALS in two independent cohorts and identified a circulating protein signature associated with survival, comprising 15 proteins associated with survival in both cohorts despite profiling on orthogonal proteomic platforms. The strong concordance of effect sizes across cohorts supports the robustness of these prognostic signals.

Several cohort-specific survival associations involved proteins previously implicated as neurodegeneration biomarkers. TNFRSF12A, which was associated with survival in both cohorts before covariate adjustment, has been implicated in presymptomatic ALS^15^, with increased levels detected before phenoconversion. PTX3^16^, ART3^17^ and IGFBP6 were identified only in the Oxford cohort, and GRN^18^, PEBP4 and IL1RAP only in the Tel Aviv cohort, reflecting differences in proteomic platforms, cohort heterogeneity, or both. Three prioritized proteins, GSN, IGFBP2 and MEGF10, showed survival associations independent of age and sex, improved prognostic accuracy in both cohorts and added prognostic value beyond clinical covariates and serum NfL, suggesting that they capture novel prognostic information.

There is supporting evidence for the involvement of GSN, IGFBP2 and MEGF10 in neurodegenerative processes. MEGF10 is a phagocytic receptor that regulates axonal debris clearance and astrocyte-mediated synapse elimination^19–21^; its levels shift early during ALS phenoconversion^15^ and are elevated in patient plasma and CSF relative to controls. GSN mutations cause hereditary amyloidosis^22^ and a rare form of bulbar-onset ALS^23^, and GSN blood levels decline in both ALS and frontotemporal dementia^24,25^. IGFBP2 is linked to Alzheimer’s disease pathogenesis, with levels rising in CSF and plasma during disease progression^26–28^. Collectively, these observations are consistent with our findings and suggest that GSN, IGFBP2 and MEGF10 capture disease-relevant biology associated with ALS progression.

The sensitivity of TNFRSF12A and other proteins to covariate adjustment suggests that they act as molecular proxies for clinical traits such as age at symptom onset or sex, reflecting the biology underlying these phenotypes.

Our findings should also be interpreted in light of differences between the proteomic platforms used in the discovery and replication cohorts. Mass spectrometry and Olink Explore HT capture partly distinct features of the circulating proteome and differ in technology and protein coverage^29^. Nonetheless, effect sizes for overlapping proteins were highly correlated across platforms, supporting the robustness of our findings and their relevance to disease biology beyond platform-specific effects.

### Limitations

Several methodological and clinical limitations should be considered. First, this study represents an initial biomarker discovery phase with exploratory value for basic and translational research rather than immediate clinical utility; translating candidate circulating markers into clinical use requires substantial further validation. Although GSN, IGFBP2 and MEGF10 are associated with subsequent survival in patients with established ALS, our findings do not demonstrate ALS specificity, diagnostic utility or the ability to distinguish ALS from disease mimics or other neurodegenerative disorders. Moreover, because the incremental predictive gain for individual-level prognosis is modest, these markers may be better suited to prognostic enrichment or risk stratification in clinical trials than to routine clinical care. Similarly, cohort-specific Kaplan–Meier tertile cut-offs cannot be used to assign an individual patient to a validated clinical risk category; dedicated assay development, standardization and calibration will be required before risk categories can be applied to new patients.

Second, although our primary prognostic proteins replicated across both cohorts, other survival associations were cohort-specific. Cross-cohort comparisons are shaped by differences in proteomic platforms and cohort composition. Although mass spectrometry quantified 2,866 circulating proteins across a broad dynamic range, it has lower sensitivity for low-abundance proteins, contributing to the lack of neurofilament light chain (NfL) detection in the Tel Aviv cohort and precluding direct comparison between NfL and our candidate proteins there, as well as limited ability to resolve post-translational modifications. Conversely, the Olink PEA used in Oxford offers greater sensitivity for low-abundance proteins. Re-profiling both cohorts on a unified platform would improve harmonization but is a major undertaking. Furthermore, the Tel Aviv and Oxford cohorts are specialized referral populations that are not fully interchangeable, differing in clinical characteristics, recruitment periods, sample handling, and survival endpoint definitions. Accordingly, cross-cohort findings are best interpreted as replication of relative biomarker associations and incremental discrimination rather than validation of identical absolute-risk estimates or calibration, which will require external validation of a fixed absolute-risk model in independent cohorts. Importantly, baseline demographic and cohort differences do not invalidate the within-cohort exposure–outcome associations observed.

Third, incomplete clinical and biochemical covariates, as well as pre-analytical factors, may affect model interpretation and generalizability. Key variables of the ENCALS survival model ^30^, including diagnostic certainty (El Escorial), frontotemporal dementia and diagnostic delay, were not uniformly available, precluding their inclusion in the primary multivariable models. Additionally, clinical laboratory and physiological measures, such as creatinine, creatine kinase, albumin, C-reactive protein, estimated glomerular filtration rate, liver enzymes, weight loss, and direct assessments of muscle mass, were unavailable in our cohorts, leaving potential residual confounding by systemic inflammation, nutritional status, renal or hepatic function, or frailty that merits mechanistic follow-up.

An additional limitation concerns the potential effects of long-term sample storage and pre-analytical handling on protein measurements. Previous studies have shown that these effects can be protein-specific and may influence measured protein abundance. Although replication across independent cohorts and proteomic platforms supports the robustness of our findings, further analytical validation, including assessments of sample stability, assay precision and inter-laboratory reproducibility, is required before clinical implementation.

## Data Availability

The data that support the findings of this study are available from the corresponding author upon reasonable request.

## Acknowledgements

E.H. is the Mondry Family Professorial Chair and head of the Andi and Larry Wolfe Center for Neuroimmunology and Neuromodulation. We thank the participants and their families for their contributions. We thank Guhan Nagappa (GSK R&D, Collegeville, PA, USA) for his support in creating the PEA/Olink database, Elham Alhathli (SITraN, University of Sheffield, UK), Sagy Krispin, Gili Wolf, and Aviad Siany (Weizmann Institute of Science) for advice and constructive conversations.

## Financial statement

This research was funded by ISF 3497/21: “PRIMALS - a precision medicine biomarkers study in ALS, based on pathology-relevant signals” - to E.H., A.A., V.D., and E.P. and DoD CDMRP grant (AL240063; GLOBALS - Global OMIC Biomarker Research for ALS). In addition, research at the Hornstein lab is funded by the Andi and Larry Wolfe Centre for Neuroimmunology and Neuromodulation, the Department of Defense – Congressionally Directed Medical Research Program, Longitude Prize on ALS by Motor Neurone Disease Association (MNDA), the Robert Packard Center for ALS Research at Johns Hopkins, the Radala Foundation for ALS Research, the ERA-Net for Research Programs on Rare Diseases (eRARE FP7) via the Israel Ministry of Health, Muscular Dystrophy Canada, the Canadian Institutes of Health Research, the Binational Science Foundation, Association Française contre les Myopathies grants 24882 and 28680, Muscular Dystrophy Association grant 1280000, Target ALS, Israel Science Foundation (ISF 3497/21, 424/22, 214/27), Minna-James-Heineman Stiftung through Minerva, Minerva Foundation with funding from the Federal German Ministry for Education and Research. Additional support was generously provided by the Weizmann - Azrieli Institute for Brain and Neural Sciences Research Centers, Weizmann - Sagol Institute for Longevity Research, Weizmann - M. Judith Ruth Institute for Preclinical Brain Research, Weizmann – Knell Family Institute for Artificial Intelligence, Kekst Family Institute for Medical Genetics, the Weizmann SABRA - Yeda-Sela-WRC Program, the Estate of Emile Mimran, The Maurice and Vivienne Wohl Biology Endowment, the Nella and Leon Benoziyo Center for Neurological Diseases, the Goldhirsh-Yellin Foundation, Dr. Sydney Brenner and Friends, the Weizmann Center for Research on

Neurodegeneration, the Redhill Foundation, the Sam and Jean Rothberg Charitable Trust, and the Dr. Dvora and Haim Teitelbaum Endowment Fund. D.G.L. receives funding from the Medical Research Council and Motor Neurone Disease Association Lady Edith Wolfson Fellowship (MR/Y001095/1, Lester/2450/795). Olink analysis was undertaken with funding from GSK Plc as part of the Institute of Molecular and Computational Medicine.

## Author contributions

H.K., I.M., and E.H. conceived the study. I.M., N.S.Y., Y.C., D.M.K., T.Z., B.A., V.D., and A.A. collected data. H.K., M.W., A.S., D.G.L., A.T., Y.Z., C.H., A.G.T., J.C.-K., and E.H. performed data analysis. A.G.S., E.P., P.J.S., A.M., M.R.T., and K.T. provided resources. H.K., M.W., A.S., J.C.-K., and E.H. wrote the manuscript. All authors reviewed and approved the final manuscript.

## Extended Data Figures

**Extended Data Fig. 1.**
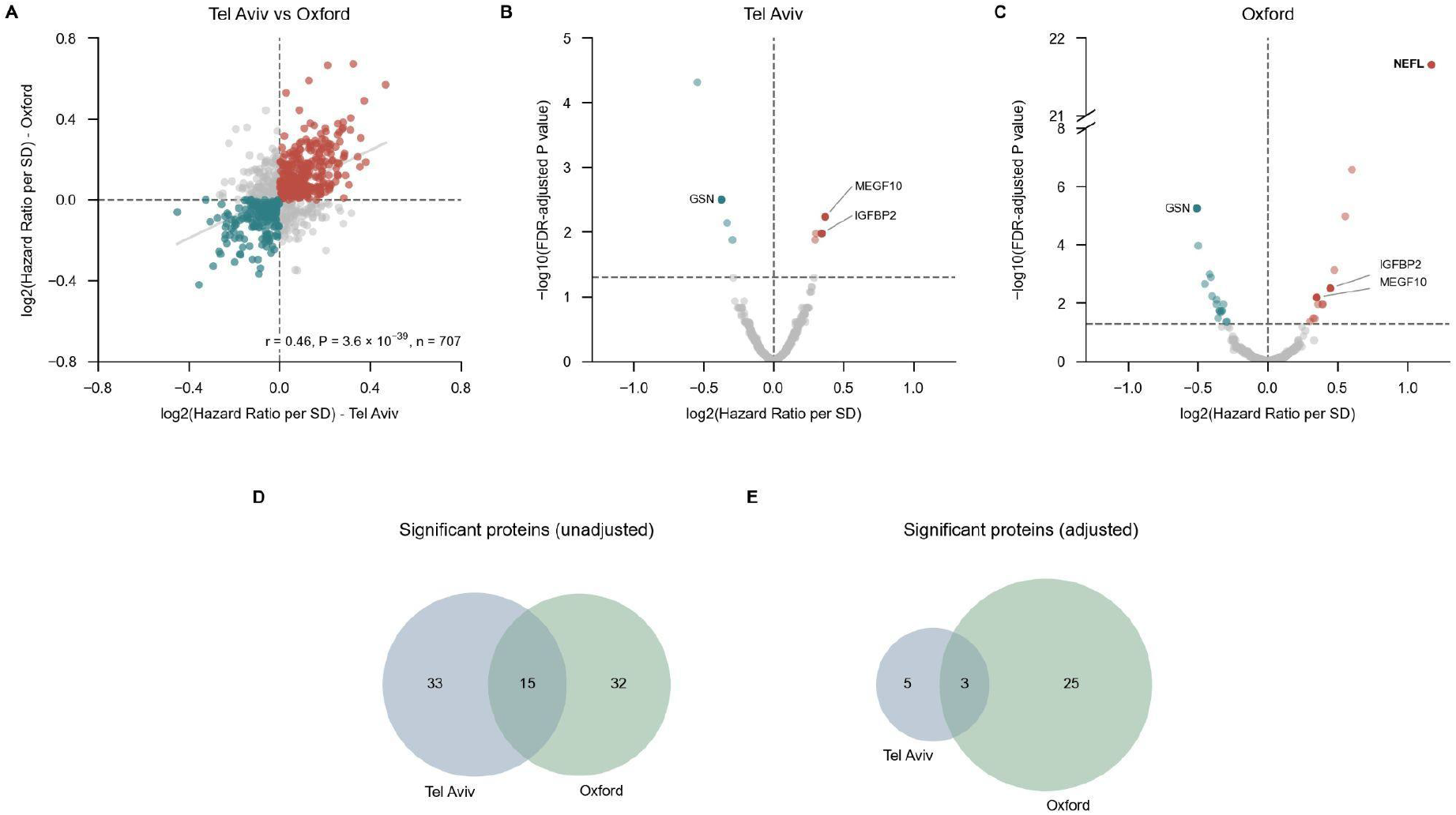
Cross-cohort comparison of survival-associated serum proteins in ALS. **a,** Cross-cohort Pearson correlation of unadjusted log2-transformed HR per s.d. Red indicates HR > 1 in both cohorts, blue indicates HR < 1 in both cohorts, and grey indicates opposite directions of association. The grey line represents the least-squares regression fit. **b,c,** Volcano plots showing associations between serum protein abundance and survival using Cox proportional hazards regression adjusted for age at symptom onset and sex in the Tel Aviv (n = 350, **b**) and Oxford (n = 380, **c)** cohorts. Log2-transformed HR (x axis) and −log10 FDR-adjusted *P* values (y axis) are shown. **d,e,** Venn diagrams showing the overlap of proteins significantly associated with survival (FDR < 0.05) between the Tel Aviv and Oxford cohorts among the 707 shared proteins, before **(d)** and after **(e)** adjustment for age at symptom onset and sex.

**Extended Data Fig. 2.**
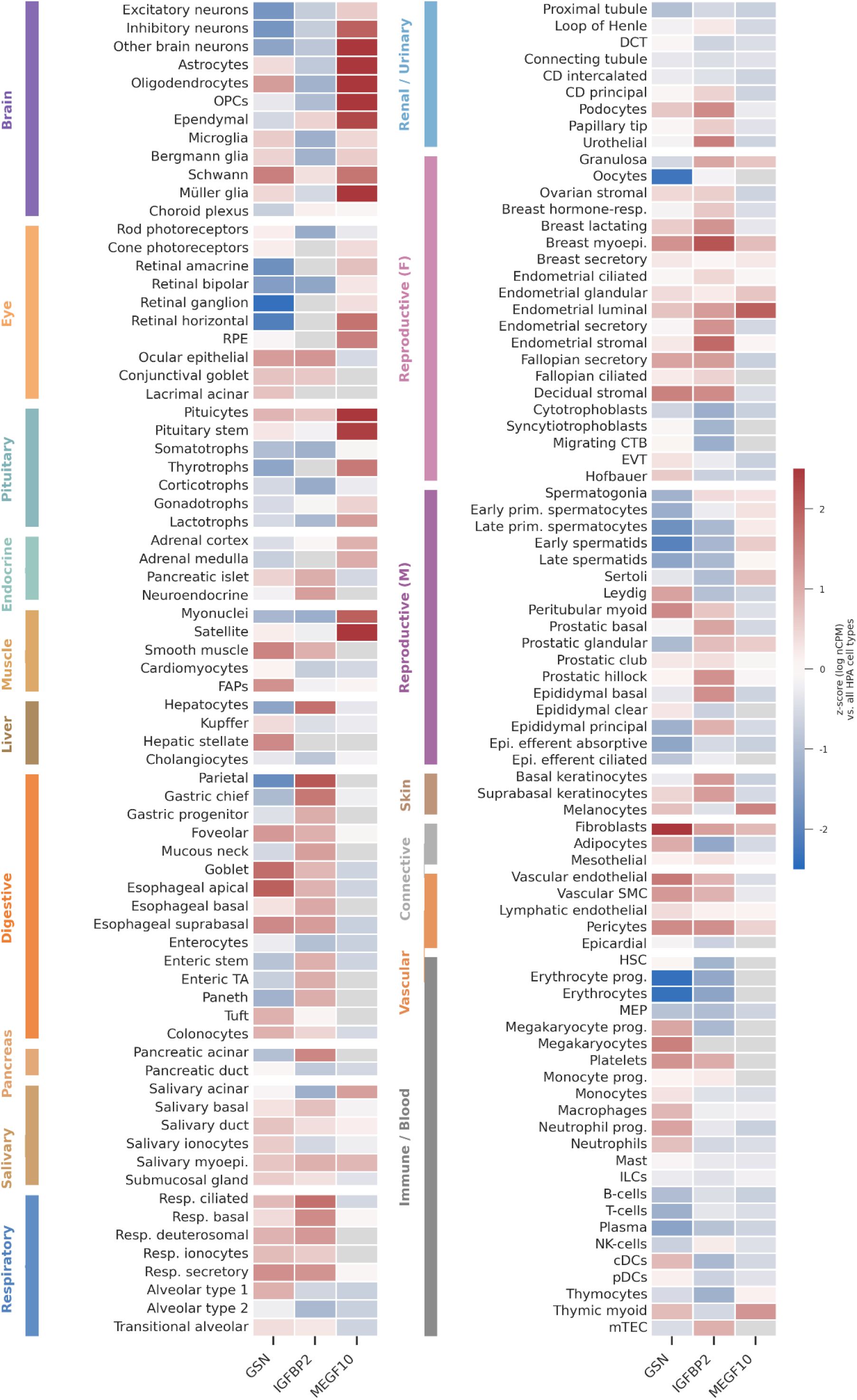
Normal-tissue transcript enrichment of candidate ALS survival-associated proteins. Heatmap of GSN, IGFBP2 and MEGF10 mRNA expression across 154 human cell types from the Human Protein Atlas single-cell transcriptomics resource. Values are log₁₀(nCPM + 1), z-scored per protein across all cell types. Colours therefore show relative enrichment within each protein, not absolute differences between proteins:red and blue indicate expression above and below that protein’s mean, and grey denotes no detectable expression (nCPM = 0). Cell types are grouped by organ system, indicated by the coloured bar at the left.

## Supplementary Figures

**Supplementary Fig. 1.**
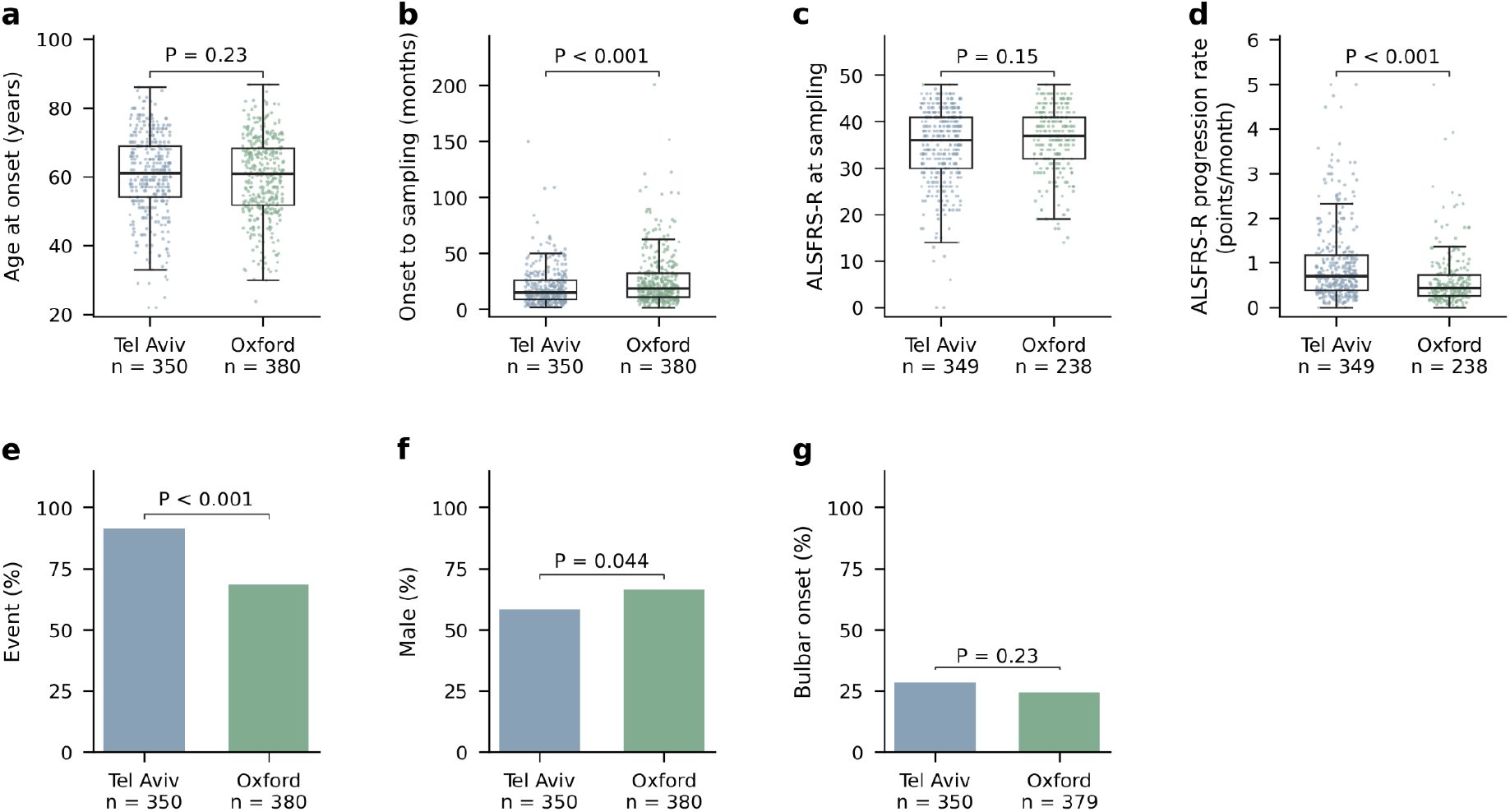
Clinical characteristics of the Tel Aviv and Oxford cohorts. **a,** Age at symptom onset. **b,** Time interval from symptom onset to sampling. **c,** ALSFRS-R score at sampling. **d,** ALSFRS-R progression rate at sampling. **e,** Proportion of deceased patients. **f,** Proportion of male patients. **g,** Proportion of patients with bulbar onset. Continuous variables **(a–d)** are presented as box plots (centre line, median; box, interquartile range; whiskers, 1.5 × interquartile range), with each dot representing an individual patient. Binary variables **(e–g)** are presented as percentages. Between-cohort comparisons were performed using two-sided Mann–Whitney *U* tests for continuous variables and χ² tests; *P* values were adjusted for multiple comparisons across all tests using the Benjamini–Hochberg procedure.

**Supplementary Fig. 2.**
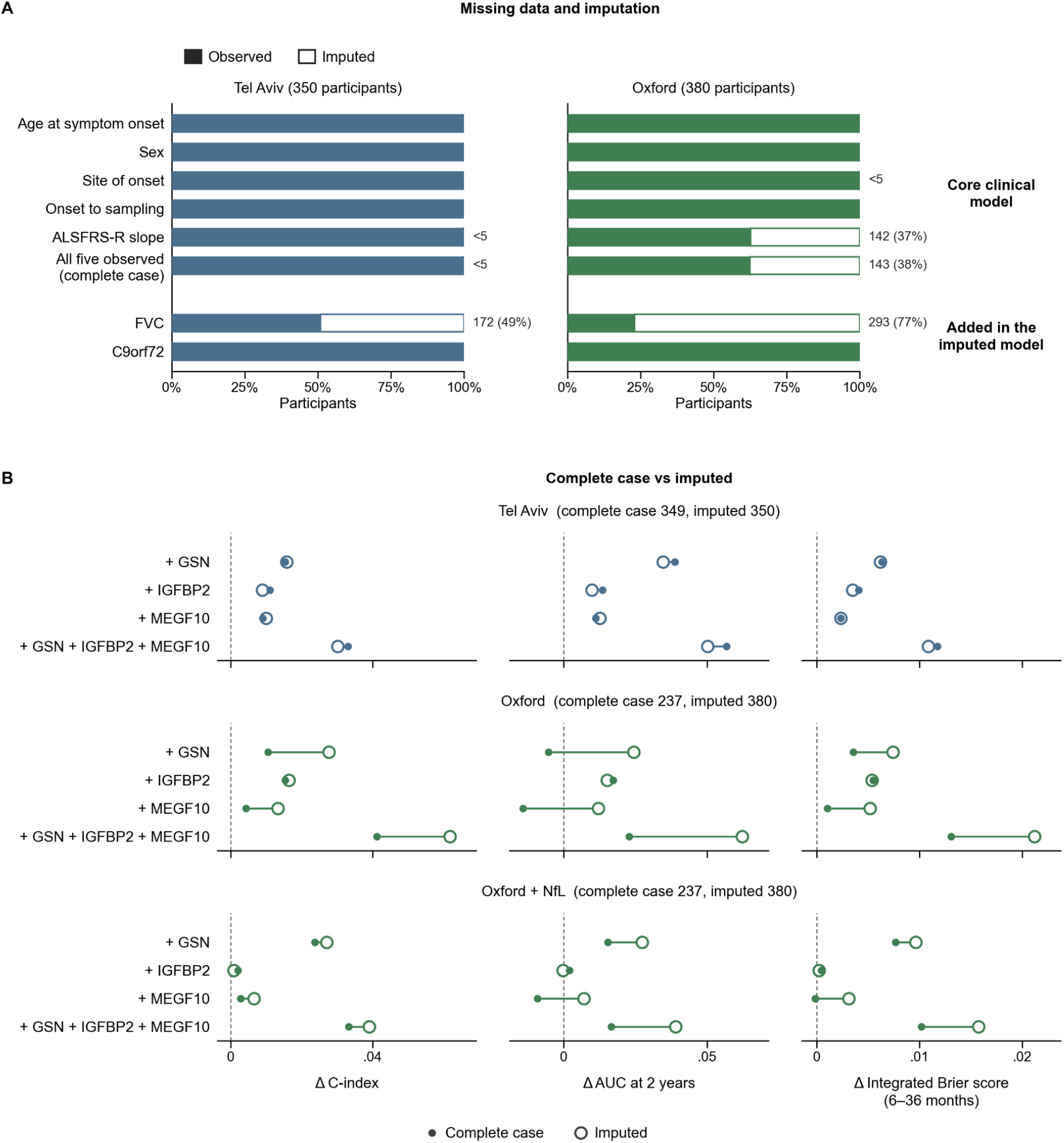
Missing clinical data and sensitivity of prediction models to imputation. **a**, Completeness of clinical covariate data in the Tel Aviv (n = 350) and Oxford (n = 380) cohorts. The upper panel shows the five core covariates included in the complete-case analysis and the proportion of participants with complete data for all five covariates. The lower panel shows FVC and C9orf72 status, which were additionally included in the clinical model using multiple imputation. Filled bars represent observed data, whereas open bars indicate missing data addressed by imputation. For the complete-case summary, the open segment represents participants missing at least one core covariate. **b,** Incremental predictive performance of GSN, IGFBP2 and MEGF10, individually or in combination, in complete-case (filled circles) versus multiply imputed (open circles) datasets, shown as gains over the clinical model for C-index, 2-year time-dependent AUC or integrated Brier score (6–36 months). Positive values indicate improvement; five repeats of 20-fold cross-validation. Full results in Extended data Table 4.

**Supplementary Fig. 3.**
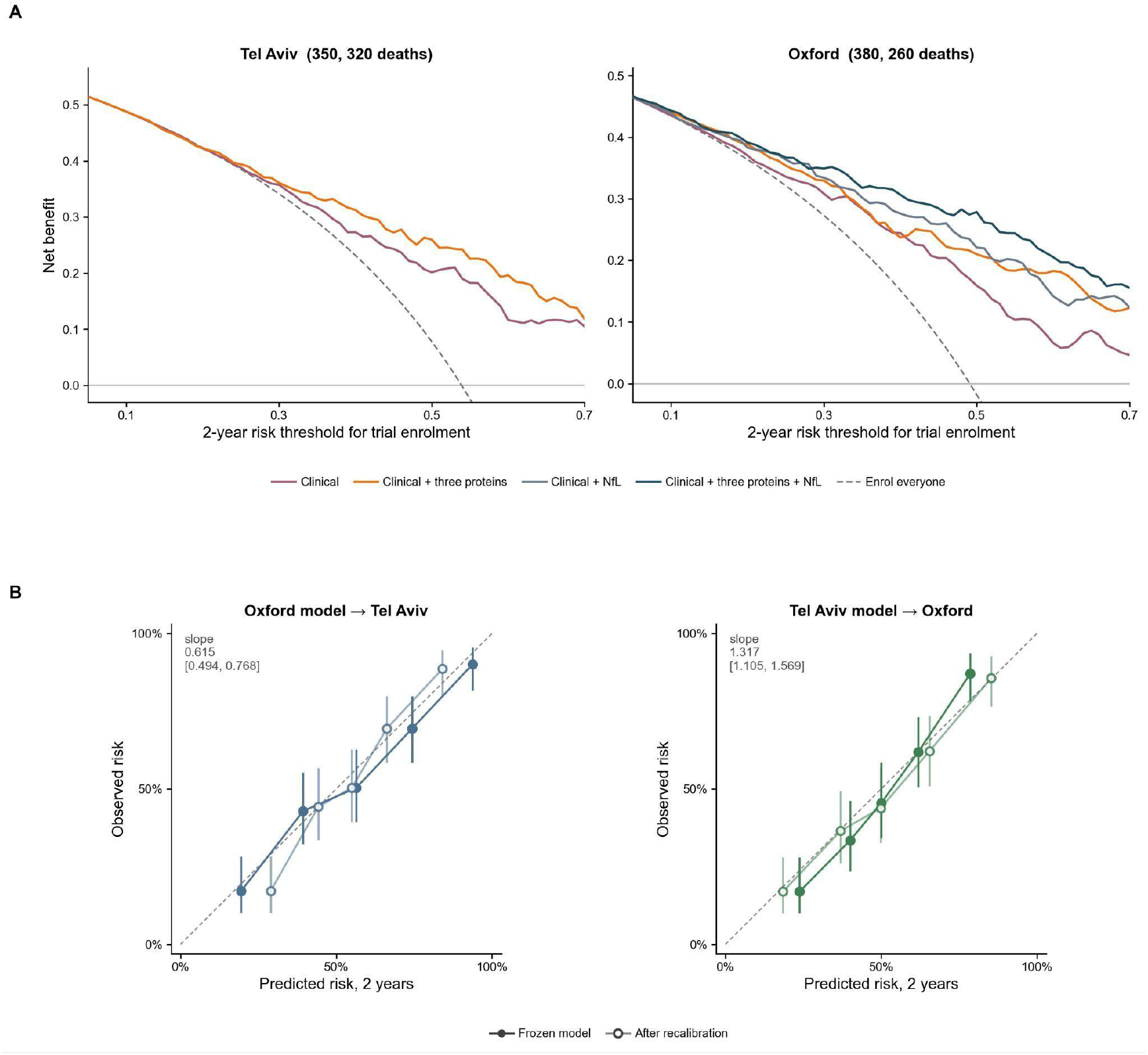
Decision-curve analysis and cross-cohort calibration of the three-protein prognostic model. The three-protein models in both panels use the same flexible parametric survival model on the hazard scale as in Fig. 4, incorporating clinical covariates, GSN, IGFBP2 and MEGF10. **a,** Decision-curve analysis at 2 years in the Tel Aviv and Oxford cohorts. Net benefit was evaluated for the clinical model and the clinical model supplemented with the three proteins, across 2-year mortality risk thresholds from 0.05 to 0.70. The dashed line represents enrolling all participants regardless of predicted risk. Models incorporating NfL are shown only for Oxford. **b,** Cross-cohort calibration at 2 years of the clinical model supplemented with the three proteins. Models were fitted in one cohort and applied to the other with coefficients held fixed: Oxford to Tel Aviv (left) and Tel Aviv to Oxford (right). Participants were divided into quintiles of predicted risk. Points show the mean predicted and observed 2-year mortality risk in each quintile of predicted risk, with 95% confidence intervals for the observed risk. Filled symbols indicate predictions from the frozen model, whereas open symbols indicate predictions after recalibration of the calibration slope in the target cohort. The dashed diagonal represents perfect calibration. The calibration slope of the frozen model and its 95% confidence interval are reported in each panel.

**Supplementary Fig. 4.**
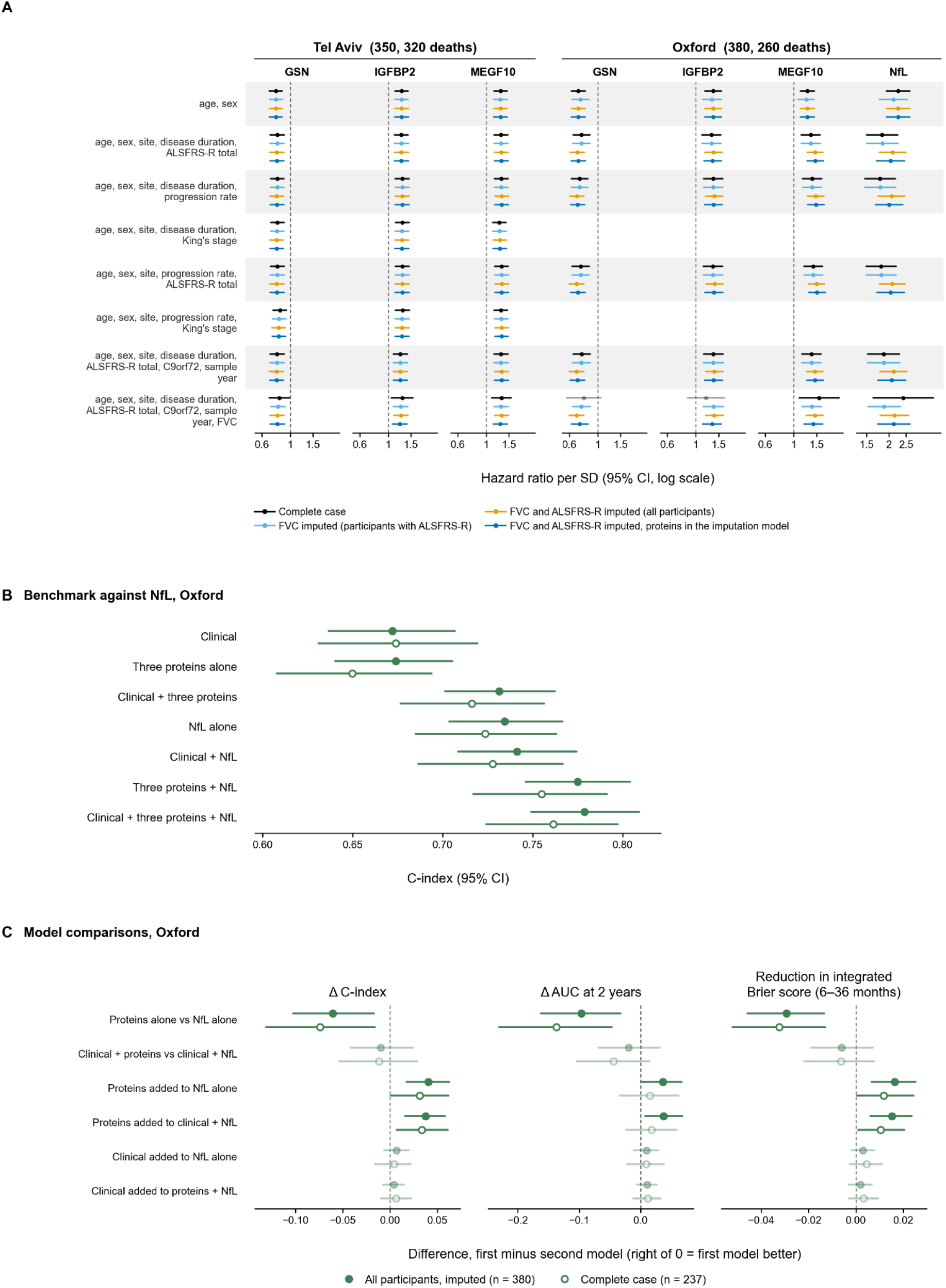
Sensitivity analyses of protein–survival associations and benchmarking of prognostic models against NfL. **a,** HRs per s.d. for GSN, IGFBP2, MEGF10 and NfL from multivariable Cox proportional-hazards models across different clinical adjustment sets in the Tel Aviv (n = 350; 320 deaths) and Oxford (n = 380; 260 deaths) cohorts. Points indicate HRs with 95% confidence intervals (CIs). Faded symbols indicate P ≥ 0.05 (two-sided Wald test). King’s stage was unavailable in Oxford. **b,** C-index (95% CI) for seven prognostic models in Oxford. Filled circles represent the imputed cohort (n = 380), and open circles represent complete cases (n = 237). The clinical model includes age, sex, site of onset, disease duration and ALSFRS-R progression rate, with FVC and C9orf72 status additionally included in the imputed analysis. **c,** Paired differences in C-index, 2-year AUC and integrated Brier score between prognostic models. Points represent differences with 95% bootstrap CIs (1,000 resamples). Positive values favour the first model; Brier score differences are sign-reversed for consistency. Faded symbols indicate P ≥ 0.05.

**Supplementary Fig. 5.**
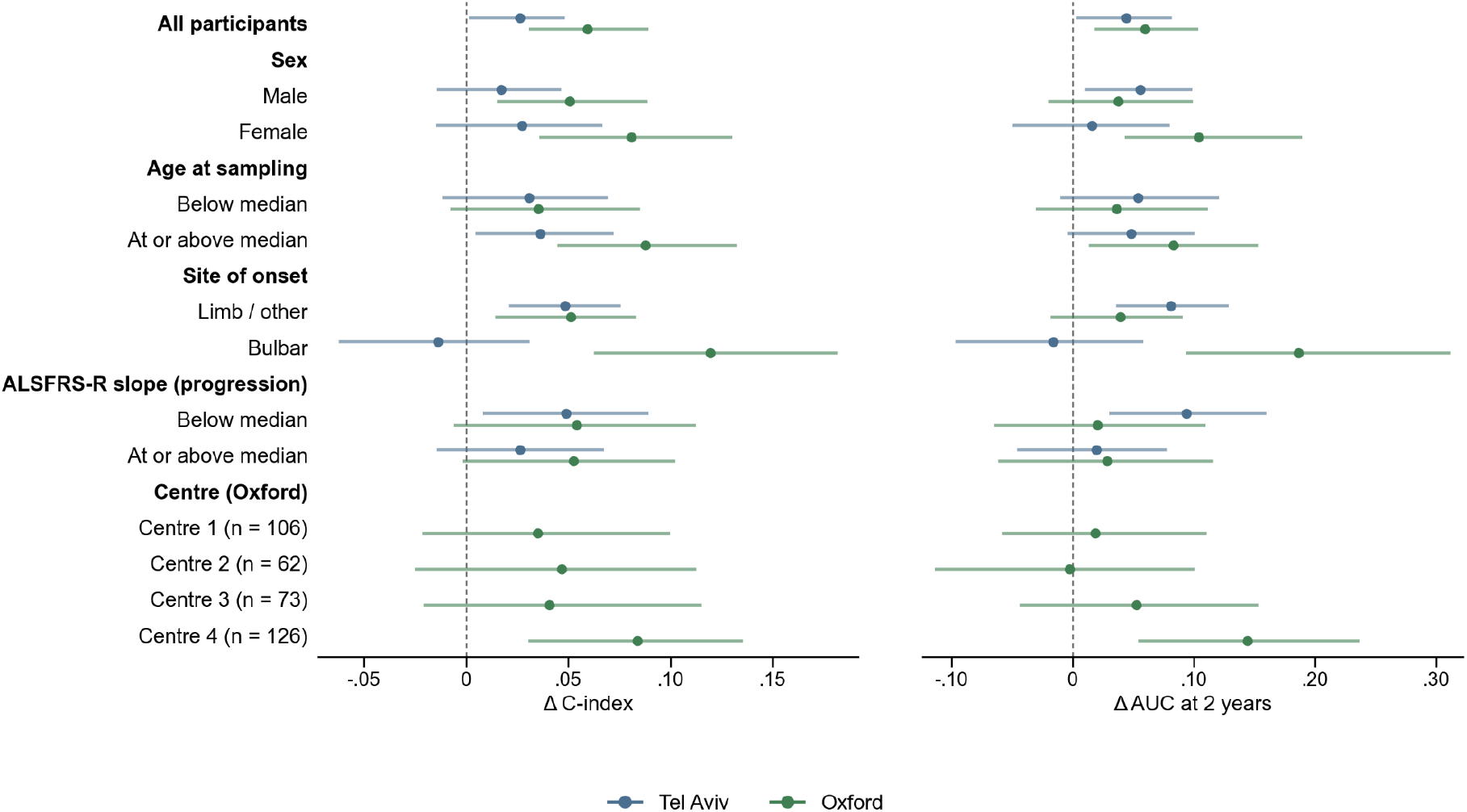
Incremental prognostic value of GSN, IGFBP2 and MEGF10 across clinical subgroups. Gain in C-index (left) and 2-year AUC (right) following the addition of the three proteins to the clinical model in the Tel Aviv (blue; n = 350) and Oxford (green; n = 380) cohorts, with missing clinical covariates addressed using multiple imputation. Points represent cross-validated estimates, and lines indicate bootstrap 95% confidence intervals. Subgroups were defined using observed values, with age and ALSFRS-R progression slope dichotomized at the cohort-specific median. Recruiting centres are shown for Oxford only, with sub-cohorts identified by numbers. Subgroup sample sizes and interaction P values are provided in Extended data Table 4.

## Methods

### Participants, ethics and study design

#### Tel Aviv cohort

An observational, retrospective cohort of patients with amyotrophic lateral sclerosis (ALS) was established at the Tel Aviv Sourasky Medical Center ALS clinic using the Gold Coast diagnostic criteria. Patients with diagnosed disease mimics were excluded. Progressive muscular atrophy (PMA) was included, whereas primary lateral sclerosis (PLS) was excluded. Frontotemporal dementia (FTD) was included only when associated with ALS. The study was approved by the Tel Aviv Sourasky Medical Center Institutional Review Board (IRB approvals 0259-22-TLV and 0245-10-TLV). Patients were enrolled between 2010 and 2021 and followed approximately every six months until death, tracheostomy, withdrawal or the end of the study, whichever occurred first. Survival time was defined in months, and riluzole treatment was documented. Survival time was defined in months, with death or tracheostomy considered an event; tracheostomy accounted for approximately 15% of the cohort. Riluzole treatment was also documented.

### Oxford cohort

An observational, retrospective cohort of patients with ALS was established at the Oxford MND Care Centre, John Radcliffe Hospital, Oxford. Participants were diagnosed by neurologists specializing in motor neuron disorders according to the Gold Coast criteria, requiring upper and lower motor neuron (UMN and LMN) dysfunction in at least one body region or LMN dysfunction in at least two regions, with investigations excluding alternative causes. Participants were recruited into six Research Ethics Committee (REC)-approved sample collection cohorts between 2009 and 2023. All participants provided informed consent, and no additional exclusion criteria were applied. The study was approved under REC references 16/LO/2136, 08/H0605/85, 14/SC/1248, 17/SC/0277, 20/WA/0027 and 21/EM/0251. Dates of death or censoring were ascertained from electronic health records in 2024.

### Clinical assessments

We followed the original ENCALS model, which used a composite survival outcome comprising death, tracheostomy, or non-invasive ventilation for more than 23 h/day as in Westeneng et al. ^30^.

### Tel Aviv cohort

Clinical data included age, sex, date and site of symptom onset (bulbar versus non-bulbar), family history of ALS and other neurodegenerative diseases, genetic testing results when available, ALS Functional Rating Scale–Revised (ALSFRS-R) scores, body mass index (BMI, kg m⁻²), forced vital capacity (FVC, percentage predicted) and manual muscle testing (MMT). Disease progression rate at enrollment was calculated as (48 − ALSFRS-R score) divided by symptom duration in months.

### Oxford cohort

Clinical assessments at sampling included date and site of symptom onset (bulbar versus non-bulbar), ALSFRS-R scores, Edinburgh Cognitive and Behavioural ALS Screen (ECAS), neurological examination, BMI and FVC (percentage predicted). Genetic testing was performed through standard clinical pathways. Disease progression rate was calculated as (48 − ALSFRS-R score) divided by symptom duration in months. Clinical UMN and LMN burden was assessed across five body regions using hyperreflexia, muscle wasting and fasciculations, yielding scores ranging from −10 to +10, representing the most flaccid to the most spastic phenotypes, respectively^31^.

### Blood collection, processing and storage

#### Tel Aviv cohort

Peripheral venous blood was collected into serum-separator tubes and centrifuged at 1,200 × g for 10 min at 4 °C. Serum supernatants were aliquoted into polypropylene tubes (Vacuette, catalog no. 455092) and stored in either vapor-phase liquid nitrogen (−180 °C) or at −80 °C within 2 h of collection. Before the present analyses, samples had undergone a maximum of two freeze–thaw cycles. Patient samples were de-identified, and laboratory personnel and data analysts were blinded to patient identities.

### Oxford cohort

Peripheral venous blood was collected into serum-separator tubes and processed according to consensus biomarker guidelines (Teunissen et al., 2009). Samples were centrifuged at 2,300 × g for 10 min at 4 °C, aliquoted into polypropylene tubes and frozen within 2 h of collection. Samples from the AMBRoSIA cohort were stored in vapor-phase liquid nitrogen (−180 °C), whereas samples from the remaining cohorts were stored at −80 °C. AMBRoSIA and PRoSNeB samples had not previously been thawed, whereas samples from the remaining cohorts had undergone one freeze–thaw cycle before the present analyses.

Storage conditions are compatible with reliable circulating biomarker measurements 32 and cryopreservation including freeze–thaw cycles maintain acceptable sample quality and do not materially affect protein measurements in line with 33–37.

### Proteomic sample preparation and profiling

#### LC–MS/MS proteomics

Serum samples (5 µL, without protease inhibitors) were depleted of the 14 most abundant proteins using High Select Top14 Resin (Thermo Scientific, #A36372), yielding ∼45–50 µg of protein per sample based on pilot measurements. Proteins were denatured and reduced in 8 M urea, 400 mM ammonium bicarbonate, and 10 mM DTT (60 °C, 30 min), then alkylated with 35 mM iodoacetamide in 100 mM ammonium bicarbonate (30 min, room temperature, dark). After diluting to 1.5 M urea and 66 mM ammonium bicarbonate, samples were digested with modified trypsin (Promega) overnight at 37 °C (1:50 w/w), followed by a second 4 h digestion (1:100 w/w). Peptides were fractionated at high pH (5–80% acetonitrile in 15 mM ammonium hydroxide, pH 10) on C18-AQ (Premier LCMS) or Oasis HLB 96-well µElution plates (Waters), concatenated non-contiguously from 20 into 3 fractions, dried, and resuspended in 0.1% formic acid [33]. Samples were processed in two batches (∼182 patients/batch; depletion/denaturation on thaw day, digestion/fractionation the following day) and analyzed at the Smoler Proteomics Center (Technion, Israel). Peptides were separated on an Evosep One system (EV1137 column, 15 cm × 150 µm; 30 SPD/44 min gradient) coupled to an Orbitrap Exploris 480 mass spectrometer (Thermo Fisher Scientific) via electrospray ionization. Positive-ion DIA data were acquired with a full MS1 scan (380–980 m/z, 60,000 resolution) and 59 MS2 windows (1 Da overlap, 3 s cycle time, 30,000 resolution, 28% HCD NCE). HeLa digests were run before and after each batch, with blanks every ∼15 injections to monitor performance and carryover.

### Olink Explore HT proteomics

Serum samples were thawed on ice and plated into 96-well plates using layouts randomized to balance biological and technical factors across and within plates^38^. Proteomic profiling was performed using Olink Explore HT (Thermo Fisher Scientific) at the Centre for Human Genetics, University of Oxford. Target-specific oligonucleotide-labeled antibody pairs underwent proximity extension assay (PEA), generating double-stranded DNA barcodes quantified via next-generation sequencing (Illumina NovaSeq) and expressed as log2-transformed Normalized Protein eXpression (NPX) values^39^. Details in ^8^.

### Proteomic data processing and quality control

#### Mass spectrometry data processing

Mass spectrometry data were searched with DIA-NN (v1.8.1) against the human UniProt reference proteome (UP000005640; May 2023 release, 81,803 entries) using default settings, with cysteine carbamidomethylation as a fixed modification, minimum peptide length of 7 amino acids, and up to 1 missed cleavage. Peptide- and protein-level false discovery rates (FDRs) were controlled at 1% via a target–decoy strategy. Label-free quantification used DIA-NN’s robust LC (high-precision) mode at the protein-group level without restricting to proteotypic peptides, followed by downstream analysis in Perseus (v1.6.15.0) ^40^. Protein intensities were summed across the three fractions per participant. No samples were excluded post-acquisition, and no further protein-level filtering (e.g., by missingness, detectability, peptide count, or coefficient of variation) was applied beyond the 1% FDR thresholds.

### Olink normalization and quality control

Assays included standard Olink internal controls (incubation, extension, detection), external sample controls for CVs, negative controls for LOD calculation (plate median plus 3 s.d.), and dilution series to prevent hook effects. Intensity normalization aligned assay medians across balanced plates. Median (Q1–Q3) intra- and inter-plate CVs were 23.7% (12.5–41.4%) and 21.2% (12.4–34.7%), respectively, with a median of 87.8% (4.9–100%) of samples below LOD per protein; sub-LOD values were retained per standard PEA practice. Six samples failing QC (>5 s.d. from the mean for PC1, PC2, mean NPX, or NPX IQR) were excluded. Covariate effects on PC1–PC10 were assessed by linear regression (*R*^2^), and PEA values were validated against paired Meso Scale Discovery (MSD) and ELISA assays^41^. Details in ^8^.

### Statistical analysis

Univariate Cox proportional hazards regression was performed for each of the 707 serum proteins measured in both the Tel Aviv discovery cohort (n = 350; 320 deaths) by data-independent acquisition mass spectrometry (DIA-MS) and the Oxford replication cohort (n = 380; 260 deaths) using the Olink Explore platform. Survival time was defined as the interval from blood sampling to death or censoring. Protein abundance was standardized within each cohort (z-score), such that HR represented the change in hazard per 1-s.d. increase in protein abundance. In each cohort, each protein was modeled separately as a continuous predictor using the CoxPHFitter function of the Python lifelines package, without penalization. Values were adjusted for multiple testing across the 707 shared proteins within each cohort using the Benjamini–Hochberg false discovery rate (FDR) procedure. Proteins with FDR < 0.05 in both cohorts were considered significantly associated with survival in both cohorts. Cross-cohort concordance of survival associations was assessed using Pearson correlation of the standardized protein effect estimates (log HRs). Univariate Cox proportional hazards models were repeated for each protein with adjustment for age at symptom onset and sex. Each model included one protein as the primary predictor, with age at symptom onset and sex included as covariates. Code used for the statistical analyses is available on GitHub at: https://github.com/HadarKlimovski/als-protein-biomarkers

### Survival prediction models

Survival was modeled using a Royston–Parmar flexible parametric survival model on the proportional hazards scale, following the architecture of the ENCALS survival model ^30,42^ as previously implemented for ALS time-to-event prediction^32(Weinreich et al), with one internal knot and a ridge penalty of 0.01 applied to all parameters except the intercept.Models were fitted by maximum likelihood using the Newton–Raphson algorithm, and the implementation was verified against the *flexsurv* R package.The parsimonious clinical reference model included age at symptom onset, sex, site of onset, time from symptom onset to sampling, and ALSFRS-R progression slope. GSN, IGFBP2 and MEGF10 were added individually and in combination to assess their incremental prognostic value. In Oxford, additional models included serum neurofilament light chain (NfL). Clinical covariates were standardized using training-set means and standard deviations, whereas protein abundances were standardized within each cohort.

Sensitivity analyses of the associations between GSN, IGFBP2, MEGF10 and survival were performed using multivariable Cox proportional hazards models with different sets of clinical covariates (Supplementary Fig. 4a).

### Nested feature selection and cross-validation

To assess the robustness of protein selection and minimize potential selection bias, we repeated the feature-selection pipeline within five repeats of 10-fold cross-validation in the Tel Aviv cohort. Within each training fold, all 1,289 proteins were screened using Cox proportional hazards models adjusted for age at symptom onset and sex, and predictive performance was evaluated on the held-out fold. Across folds, PEBP4 was selected in 98% of cases, followed by GSN (42%), MEGF10 (18%) and IGFBP2 (4%). This approach ensured that feature selection was performed independently of the held-out outcomes.

### Selection-frequency threshold

To distinguish stable protein selection from that expected by chance, we established cohort-specific selection-frequency thresholds using permutation analysis. For each cohort, we generated 1,000 null datasets by randomly permuting participants’ complete protein profiles relative to their survival outcomes, age and sex. This preserved correlations among proteins and the associations between clinical covariates and survival while disrupting protein–survival associations. For each permuted dataset, we repeated the original bootstrap selection procedure and calculated the selection frequency of every protein. We then recorded the highest selection frequency across all proteins within each permutation, yielding a null distribution of 1,000 maximum selection frequencies per cohort. The selection threshold was defined as the 95th percentile of this distribution, such that the probability of at least one protein exceeding the threshold by chance was approximately 5% under the permutation null. A single threshold was then applied to all proteins within each cohort. The resulting thresholds were 83.0% for Tel Aviv and 76.0% for Oxford, corresponding to an empirical family-wise error rate of approximately 5% under the permutation null.

### Missing data and multiple imputation

Missing clinical covariates were multiply imputed separately within each cohort using predictive mean matching implemented in the *mice* R package (v3.19.0; 10 imputations, 10 iterations). FVC was missing in 49.1% of Tel Aviv and 77.1% of Oxford participants, whereas ALSFRS-R progression slope was missing in 0.3% and 37.4%, respectively. Bulbar onset was additionally imputed for one participant in Oxford. Missing C9orf72 status was not imputed; participants without a recorded genetic test result (288 of 350 in Tel Aviv and 287 of 380 in Oxford) were classified as non-carriers. Complete-case prediction analyses were also performed as sensitivity analyses in participants with complete data for the five core clinical covariates (n = 349 in Tel Aviv and n = 237 in Oxford), excluding FVC and C9orf72 status.To assess potential selection bias, we compared the clinical characteristics and survival of participants included in and excluded from the complete-case analysis (Extended Data Table 4 and Supplementary Fig. 2).

### Internal validation and predictive performance

Predictive performance was evaluated using five repeats of 20-fold cross-validation. In each repeat, participants were randomly divided into 20 folds. Models were trained on 19 folds and used to predict risk scores for participants in the remaining fold, until every participant had received one out-of-fold prediction. This procedure was repeated five times using new random fold assignments, with identical folds maintained across models within each cohort. Cross-validation was performed separately within each of the ten imputed datasets. For each participant, the resulting risk scores were averaged across repeats and imputations to obtain a single final risk score. The C-index was then calculated once for each model using these averaged risk scores across all participants, rather than averaging C-indices calculated separately for individual folds. Discrimination was assessed using Harrell’s concordance index (C-index) and Uno’s cumulative/dynamic time-dependent area under the receiver operating characteristic curve (AUC). Prediction accuracy was assessed using the inverse probability of censoring-weighted (IPCW) Brier score at 6, 12, 18, 24, 30 and 36 months, and the integrated Brier score over 6–36 months. Incremental predictive performance was quantified as the difference between the protein-augmented and reference models, with Brier score differences calculated as reference minus augmented model so that positive values indicated improvement for all metrics. Percentile 95% confidence intervals (CIs) were estimated from 1,000 participant-level bootstrap resamples of the averaged out-of-fold predictions. Paired differences between models were calculated using identical resamples, with predictions held fixed. The incremental predictive contribution of individual proteins was assessed using 1,000 permutations of protein abundances within each training fold, followed by model refitting and cross-validation. Empirical *P* values were calculated as (1 + the number of permutations yielding performance gains at least as large as the observed gain)/1,001. For single-protein models, the corresponding protein was permuted. For the three-protein model, each protein was permuted separately while retaining the other two, and the least significant result was reported for each performance metric. In comparisons against the clinical model including NfL, NfL was retained and only the candidate proteins were permuted. Complete-case and imputed performance comparisons were based on mean cross-validated performance.

To assess whether the incremental prognostic value of GSN, IGFBP2 and MEGF10 varied across clinical subgroups, we evaluated gains in C-index and 2-year AUC within each subgroup. Age at symptom onset and ALSFRS-R progression slope were dichotomized at cohort-specific medians, and recruiting sub-cohorts were examined in Oxford only. Subgroup-specific performance gains were estimated using cross-validated predictions, with 95% confidence intervals obtained by bootstrap resampling (Supplementary Fig. 5; Extended Data Table 4).

### Cross-cohort validation

Models were trained independently within each cohort and evaluated externally in the other without refitting. All model coefficients, baseline survival functions, clinical covariate standardization parameters, and risk-group cut-offs were fixed from the training cohort. To account for cross-platform scaling differences between DIA-MS and Olink PEA, protein abundances were standardized within the validation cohort without reference to survival outcomes. Confidence intervals and paired model comparisons were derived from 1,000 bootstrap resamples, repeating validation-cohort protein standardization within each resample.

### Calibration and risk stratification

Calibration was assessed both within each cohort and externally. Within each cohort, calibration was evaluated using out-of-fold cross-validated predictions. We calculated calibration slopes using the flexible parametric survival model, with Cox-based calibration slopes also reported, calibration-in-the-large at 24 months, and the median absolute error of predicted survival among participants who died. Calibration curves at 24 months were constructed by comparing mean predicted mortality risk within deciles of predicted risk with observed mortality estimated using the Kaplan–Meier method. Bootstrap 95% confidence intervals (CIs) were calculated for all calibration measures (Extended data table Table 4).

For external calibration, the model fitted in one cohort was frozen and applied without refitting to the other cohort, in both directions. Calibration was evaluated at 24 months by comparing mean predicted mortality risk within quintiles of predicted risk with observed mortality estimated using the Kaplan–Meier method. Calibration slopes and calibration-in-the-large, with 95% CIs, were calculated. Recalibration was performed in the target cohort by retaining the original linear predictor and re-estimating baseline survival and the calibration slope using three repeats of 10-fold cross-validation.

For risk stratification, Oxford participants were classified using either the tertile cut-offs of the linear predictor derived in Tel Aviv or fixed thresholds of predicted 2-year mortality risk (<40%, 40–70% and >70%). Survival differences between groups were assessed using log-rank tests, and hazard ratios (HRs) comparing the highest- and lowest-risk groups were estimated using Cox proportional hazards regression.

### Gene set enrichment analysis

Protein–survival associations were modeled separately in the Tel Aviv and Oxford cohorts via Cox proportional hazards regression adjusted for age at symptom onset and sex. All proteins were ranked without significance filtering by signed Wald Z statistics (β/*SE*), and pre-ranked GSEA was run using fgsea (v1.38.0; multilevel method, nPermSimple = 5,000) across Reactome and Gene Ontology Biological Process gene sets (10–500 genes) retrieved via msigdbr (v26.1.1). The same pipeline was applied to the independent Sheffield Olink cohort. Immune pathways were defined by predefined immune and inflammatory keywords. Proportions of positive normalized enrichment scores (NES) in immune versus non-immune pathways were compared using two-sided Fisher’s exact tests with Woolf 95% confidence intervals for odds ratios. Cross-cohort NES correlation was evaluated by Pearson correlation, and pathways with nominal *P* < 0. 05 and matching NES signs in both Tel Aviv and Oxford were deemed nominally replicated. Redundant pathways were pruned at leading-edge overlap of 80% or greater or Jaccard similarity of 0.70 or greater, leaving 20 representative pathways for visualization.

### Pre-diagnosis plasma proteomic analysis in UK Biobank

Pre-diagnosis plasma proteomic data (Olink Explore) were obtained from UK Biobank for motor neuron disease (MND; ICD-10 G12.2), Alzheimer’s disease (G30.1), all-cause dementia (F00), and Parkinson’s disease (G20), ascertained from inpatient records. Time to diagnosis was defined from recruitment blood draw to the first inpatient ICD-10 record; post-diagnosis samples were excluded, leaving 259 of 281 MND cases. Because each participant provided one baseline sample, analyses reflect cross-sectional variation across pre-diagnosis intervals rather than intra-individual trajectories. Across all 2,969 proteins (with NEFL as a positive control), abundance was modeled against time to diagnosis using generalized cross-validated smoothing splines (*smooth.spline*, 4,000-point grid). Changepoints were identified at the maximum perpendicular distance from the chord connecting the curve endpoints, and Pearson correlations with time to diagnosis were computed on either side (negative *r* indicating higher levels nearer diagnosis), with Benjamini–Hochberg FDR correction. For case–control analyses, each disease cohort was stratified into pre-diagnosis time tertiles prior to inspecting proteomic data (MND cut-points: 1,524 and 2,516 days). Each tertile was matched 1:5 to unique healthy controls without replacement (exact sex match, nearest-neighbor recruitment age within a 3-year caliper; greedy matching, seed 20260924). Complete-case linear models adjusted for age and sex estimated case–control differences in control s.d. units, with Benjamini–Hochberg adjustment across 2,974 proteins per tertile.

### Split-slope analysis of time before diagnosis

For each protein a smoothing spline was fitted to plasma level against days before diagnosis following ^42^ and predicted on a 4,000-point grid. The changepoint was taken as the grid point of maximum perpendicular distance from the chord joining the first and last grid points, and a Pearson correlation was computed on each side of it. As days before diagnosis increases away from diagnosis, a negative correlation indicates a protein rising as diagnosis approaches. The procedure was applied unchanged to all 2,969 assayed proteins, and P values were corrected across that panel by the Benjamini-Hochberg method. NEFL was analysed as a positive control.

### Matched case-control differential abundance

Cases were the same 259 pre-diagnosis participants, split into tertiles of days before diagnosis at 1,524 and 2,516 days. Tertiles were fixed before any protein result was examined. Each tertile was matched separately to healthy controls drawn from the 4,567 participants carrying the UK Biobank healthy-control flag and no MND ICD10 code: exact on sex, nearest age at recruitment, five controls per case, without replacement, with a caliper of three years. Controls were not reused between tertiles. For each of 2,974 proteins we fitted a linear model of plasma level on case status with age and sex as covariates, taking the case-status coefficient as the effect estimate and scaling it by the standard deviation of the matched control group.

### Cross-cohort direction concordance

Survival effects in the Tel Aviv cohort were taken from a Cox model of survival from blood draw on plasma protein level with age and sex as covariates, restricted to the 1,218 proteins retained by the provider’s quality control. Because that model was fitted on raw protein units, each coefficient was rescaled to one standard deviation of its own protein, computed on the same complete-case rows, which is exact for a Cox model and requires no refitting. These were intersected with the UK Biobank split-slope results, giving 607 proteins measured in both, and ranked

### Cell-type origin analysis

To determine the cellular origin of the candidate serum proteins, we used the Human Protein Atlas single-cell transcriptomics resource^43,44^. Normalized expression values (nCPM) were obtained for 154 curated cell types across major organ systems. Undifferentiated and differentiating spermatogonia were combined into a single category, resulting in 153 cell types. Expression values were log10-transformed [log10(nCPM + 1)] and standardized within each gene using row-wise z-scores to visualize relative expression across cell types. Cell types with zero expression were masked in grey. Heatmaps were displayed using a diverging color scale centered at zero and clipped at ±2.5. Cell types were grouped by organ system and displayed in two panels. Absolute nCPM values and corresponding z-scores are provided in Extended data Table 2.

